# Socioeconomic position and SARS-CoV-2 infections: seroepidemiological findings from a German nationwide dynamic cohort

**DOI:** 10.1101/2021.06.21.21259001

**Authors:** Jens Hoebel, Markus M. Grabka, Carsten Schröder, Sebastian Haller, Hannelore Neuhauser, Benjamin Wachtler, Lars Schaade, Stefan Liebig, Claudia Hövener, Sabine Zinn

**Affiliations:** Robert Koch Institute, Department of Epidemiology and Health Monitoring, Berlin, Germany; German Institute for Economic Research, Socio-Economic Panel, Berlin, Germany; Freie Universität Berlin, School of Business and Economics, Berlin, Germany; Robert Koch Institute, Department of Infectious Disease Epidemiology, Berlin, Germany; Robert Koch Institute, Centre for Biological Threats and Special Pathogens, Berlin, Germany; Humboldt University, Departement of Social Sciences, Berlin, Germany

## Abstract

**Background:** Evidence on the relationship between socioeconomic position (SEP) and infections with the SARS-CoV-2 coronavirus is still limited as most of the available studies are ecological in nature. This is the first German nationwide study to examine differences in the risk of SARS-CoV-2 infections according to SEP at the individual level.

**Methods:** The ‘CORONA-MONITORING bundesweit’ (RKI-SOEP) study is a seroepidemiological survey among a dynamic cohort of the German adult population (n=15,122; October 2020 – February 2021). Dried blood samples were tested for SARS-CoV-2 antibodies and oral-nasal swabs for viral RNA. SEP was measured by education and income. Robust logistic regression was used to examine adjusted associations of SARS-CoV-2 infections with SEP.

**Results:** 288 participants were seropositive, PCR-positive, or self-reported a previous laboratory-confirmed SARS-CoV-2 infection. The adjusted odds of SARS-CoV-2 infection were 1.87-fold (95% confidence interval [CI]=1.06-3.29) higher among low-educated than highly educated adults. Evidence was weaker for income differences in infections (odds ratio=1.65; 95% CI=0.89-3.05). Highly educated adults had lower odds of undetected infection.

**Conclusion:** The results indicate an increased risk of SARS-CoV-2 infection in low-educated groups. To promote health equity in the pandemic and beyond, social determinants should be addressed more in infection protection and pandemic planning.

## INTRODUCTION

The SARS-CoV-2 (Severe Acute Respiratory Syndrome Coronavirus 2) virus has spread rapidly worldwide. In Germany, the first cases were identified in the federal state of Bavaria at the end of January 2020 [1]. By early March 2020, infections with the virus were reported from all German federal states. Initial research on the pandemic indicates that SARS-CoV-2 infections occur more frequently in socioeconomically deprived areas [2]. Infection risks may accordingly be higher for people in disadvantaged living and certain working conditions, especially during more advanced stages of the pandemic [3].

In the occupational context, for example, essential workers employed in the logistic, healthcare, retailing or public transport sector, who tend to have lower incomes than non-essential workers [4], often work in conditions involving closer physical proximity to others. The possibility to work remotely is, by contrast, generally more available to people with higher incomes and qualifications [5]. Furthermore, crowded living conditions and limited access to effective personal protective equipment may increase the risk of viral transmissions and thereby produce inequalities in infections [6].

To date, research on socioeconomic differences in SARS-CoV-2 infections has been based mostly on ecological studies, which have correlated area-level rates of infections with area-level socioeconomic indicators [2]. The advantage of ecological studies is that area-level data are available relatively quickly, for example from existing surveillance systems. Ecological studies can thus be an expedient starting point for exploring the phenomenon. However, their findings are prone to ecological fallacy, areas can be very heterogeneous in terms of their residents’ SEP and inequalities are probably underestimated. Seroepidemiological individual-level studies are therefore needed to examine the relationship between socioeconomic position (SEP) and SARS-CoV-2. In this study, we introduced serological and polymerase chain reaction (PCR) testing along with a SARS-CoV-2-specific questionnaire into an existing German cohort and investigated whether SARS-CoV-2 infections were associated with SEP at the individual level.

## METHODS

### Study design and data

The ‘CORONA-MONITORING bundesweit’ study (RKI-SOEP study) is based on the nationwide population-based random samples administered by the German Socio-Economic Panel (SOEP), a dynamic cohort from Germany’s resident population in private households [7]. A gross sample of 31,675 adult cohort members was invited to participate, of which 15,122 individuals (response: 48%) participated in the study (*Figure 1*).

**Figure 1.**
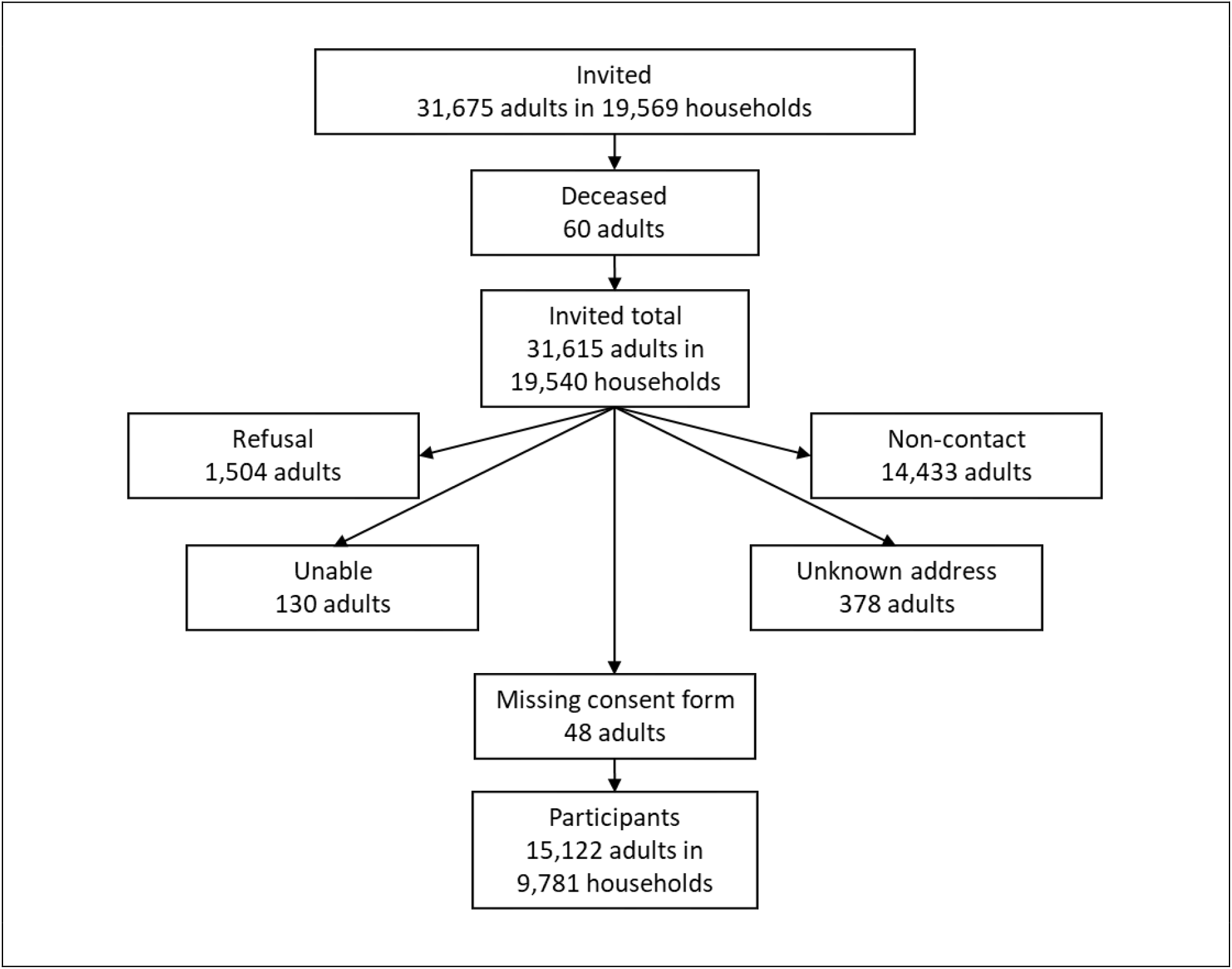
Flowchart of participation in the RKI-SOEP study, Germany, October 2020 – February 2021

Between October 2020 and February 2021, biospecimens and interview data were collected once from each participant. Participants provided a dried capillary blood sample obtained by finger prick to serologically detect antibodies from a previous SARS-CoV-2 infection by Enzyme-linked Immunosorbent Assay (ELISA). To test for a current infection, participants provided an oral-nasal swab sample for PCR testing. Both specimens were collected by the participants themselves using CE-certified sample collection and submission kits sent by post along with written, pictorial, and video instructions. Interview data were collected using a one-page paper questionnaire including questions on previous throat swab laboratory tests for SARS-CoV-2. Further details can be found in the study protocol [7].

### Measures and definitions

We used three metrics of SARS-CoV-2 infection based on combinations of the following: seropositivity for SARS-CoV-2 IgG antibodies from dried blood samples (Euroimmun SARS-CoV-2-S1 IgG-ELISA; cut-off adapted for dried blood spot testing: ratio ≥ 0.94 [8]), a positive SARS-CoV-2-PCR test result in the study, and any self-reported SARS-CoV-2-positive throat swab laboratory test conducted prior to study participation. Cases were defined as those meeting at least one of these criteria. Undetected infection with SARS-CoV-2 was defined as having tested seropositive or PCR-positive during the study but self-reported never having had a SARS-CoV-2-positive swab test before.

SEP was measured using participants’ self-reported pre-pandemic (and hence exogenous) education and income from SOEP wave 2019 or the latest available from earlier waves. Using the CASMIN educational classification, participants’ highest school and vocational qualifications were classified into low (no, primary or low secondary education), middle (intermediate/high secondary education) and high (tertiary education). Equivalised current disposable household income was calculated using the OECD-modified equivalence scale and categorised into low (<60% of median), middle (60% - <150% of median), and high (≥150% of median).

### Statistical analysis

The prevalence of SARS-CoV-2 infections was estimated using weighting factors to compensate for sampling design and non-random non-response. The weighting factors result from complex non-response modelling at the person and household level and calibration of the sample to match the official German population statistics by age, sex, federal state, municipality size, household size and owner-occupied housing. Standard error estimation was performed using Stata’s survey data commands (version 17.0, StataCorp LLC, College Station, TX, USA) accounting for weighting and household clustering. Logistic regression models with household-clustered standard errors were used to estimate odds ratios (OR) for SARS-CoV-2 infections by SEP, adjusted for a set of covariates to control for potential confounding: age, sex, household size, migrant background, urban–rural residence, region (east/west) and date of participation.

## RESULTS

*Table 1* presents the sociodemographic characteristics of the study population and shows that more than 80% of all subjects participated in October and November 2020 (median participation date: 11 November 2020). Among the 15,122 study participants aged 18 to 99 years, 192 were seropositive for SARS-CoV-2 IgG antibodies, 51 were PCR-positive for SARS-CoV-2 RNA, and 146 self-reported having had a laboratory-confirmed SARS-CoV-2 infection before study participation. At least one of these criteria was met by 288 participants.

**Table 1.** Characteristics of the study population (n=15,122)

|  | n | %<sup>a</sup> |
| --- | --- | --- |
| **Sex** |  |  |
| Women | 8,099 | 50.9 |
| Men | 7,023 | 49.1 |
| **Age group** |  |  |
| 18–34 years | 2,805 | 25.0 |
| 35–49 years | 3,553 | 22.7 |
| 50–64 years | 4,945 | 27.6 |
| 65–79 years | 3,126 | 18.4 |
| 80+ years | 693 | 6.4 |
| **Education** |  |  |
| Low | 3,267 | 31.0 |
| Middle | 6,326 | 41.9 |
| High | 4,916 | 27.1 |
| Missing | 613 | – |
| **Income** |  |  |
| <60% of median | 1,524 | 15.2 |
| 60% – <150% of median | 8,406 | 57.6 |
| ≥150% of median | 5,065 | 27.2 |
| Missing | 127 | – |
| **Date of participation** |  |  |
| October 2020 | 5,532 | (36.6) |
| November 2020 | 6,748 | (44.6) |
| December 2020 | 2,090 | (13.8) |
| January 2021 | 595 | (3.9) |
| February 2021 | 157 | (1.0) |
| **SARS-CoV-2 IgG antibodies** |  |  |
| Seropositive | 192 | 1.3 |
| Seronegative | 14,589 | 98.7 |
| Missing | 341 | – |
| **SARS-CoV-2 RNA** |  |  |
| PCR-positive | 51 | 0.4 |
| PCR-negative | 14,638 | 99.6 |
| Missing | 433 | – |
| **Previously tested positive for SARS-CoV-2<sup>b</sup>** |  |  |
| Yes | 146 | 1.1 |
| No | 14,771 | 98.9 |
| Missing | 205 | – |

<sup>a</sup> weighted percentage (unweighted percentage in brackets)<sup>b</sup> self-reported positive throat swab test before study participation

*Table 2* shows the prevalence and adjusted odds ratios for SARS-CoV-2 infection by SEP. Seropositivity, sero-or PCR-positivity, and a self-reported positive swab test were each most prevalent in the lowest education and income groups. After adjusting for covariates, low education remained associated with seropositivity and the combined infection indicator of measured sero-or PCR-positivity. A known — i.e. previously detected — infection with SARS-Co-V-2 as indicated by a self-reported positive swab test prior to the study showed no consistent association with either education or income. When all three infection parameters were combined in one infection indicator (sero-/PCR-positive or previously tested positive), the odds ratio for SARS-CoV-2 infection was 1.87-fold higher in the low than high education group. Evidence was weaker for income differences in infections. With regard to undetected infections, highly educated adults had lower odds of being sero-or PCR-positive without previously having received a positive swab test result compared to those with middle or low education, net of all covariates (adjusted OR comparing high vs. low/middle education: 0.45, 95% confidence interval 0.22-0.93, *p*=0.031).

**Table 2.** SARS-CoV-2 infection by socioeconomic position among adults in Germany, October 2020 – February 2021

|  | % (95% CI)<sup>a</sup> | OR (95% CI)<sup>b</sup> | *p* |
| --- | --- | --- | --- |
| **Seropositive** |  |  |  |
| **Education** |  |  |  |
| Low | 1.80 (1.19–2.71) | 2.32 (1.18–4.53) | 0.014 |
| Middle | 0.98 (0.67–1.43) | 1.20 (0.69–2.08) | 0.527 |
| High | 1.01 (0.69–1.50) | Ref. |  |
| **Income** |  |  |  |
| <60% of median | 2.04 (1.12–3.70) | 1.56 (0.77–3.17) | 0.220 |
| 60% – <150% of median | 1.20 (0.87–1.66) | 1.02 (0.58–1.79) | 0.945 |
| ≥150% of median | 1.02 (0.69–1.52) | Ref. |  |
| **Sero-/PCR-positive** |  |  |  |
| **Education** |  |  |  |
| Low | 2.05 (1.37–3.07) | 2.03 (1.10–3.75) | 0.024 |
| Middle | 1.46 (1.04–2.05) | 1.45 (0.89–2.37) | 0.140 |
| High | 1.21 (0.85–1.72) | Ref. |  |
| **Income** |  |  |  |
| <60% of median | 2.69 (1.29–5.51) | 1.58 (0.77–3.26) | 0.213 |
| 60% – <150% of median | 1.53 (1.15–2.04) | 1.09 (0.65–1.82) | 0.746 |
| ≥150% of median | 1.18 (0.82–1.70) | Ref. |  |
| **Previously tested positive<sup>c</sup>** |  |  |  |
| **Education** |  |  |  |
| Low | 1.39 (0.82–2.35) | 1.68 (0.76–3.69) | 0.197 |
| Middle | 0.84 (0.54–1.29) | 0.87 (0.43–1.76) | 0.694 |
| High | 1.17 (0.74–1.86) | Ref. |  |
| **Income** |  |  |  |
| <60% of median | 1.99 (0.94–4.17) | 1.42 (0.61–3.30) | 0.418 |
| 60% – <150% of median | 0.98 (0.68–1.42) | 0.82 (0.40–1.69) | 0.596 |
| ≥150% of median | 1.06 (0.59–1.88) | Ref. |  |
| **Sero-/PCR-positive or previously tested positive** |  |  |  |
| **Education** |  |  |  |
| Low | 2.44 (1.71–3.48) | 1.87 (1.06–3.29) | 0.029 |
| Middle | 1.78 (1.31–2.42) | 1.29 (0.78–2.14) | 0.315 |
| High | 1.68 (1.18–2.39) | Ref. |  |
| **Income** |  |  |  |
| <60% of median | 3.64 (2.10–6.24) | 1.65 (0.89–3.05) | 0.112 |
| 60% – <150% of median | 1.81 (1.39–2.33) | 0.95 (0.57–1.56) | 0.827 |
| ≥150% of median | 1.63 (1.09–2.43) | Ref. |  |
| **Undetected infection** |  |  |  |
| **Education** |  |  |  |
| Low | 1.00 (0.60–1.66) | 2.17 (0.90–5.25) | 0.085 |
| Middle | 0.91 (0.58–1.43) | 2.27 (1.03–5.00) | 0.042 |
| High | 0.51 (0.29–0.89) | Ref. |  |
| **Income** |  |  |  |
| <60% of median | 1.56 (0.75–3.24) | 1.77 (0.81–3.83) | 0.150 |
| 60% – <150% of median | 0.77 (0.52–1.13) | 1.03 (0.54–1.96) | 0.855 |
| ≥150% of median | 0.64 (0.41–1.00) | Ref. |  |

<sup>a</sup> prevalence (weighted); OR, odds ratio; CI, confidence interval; Ref., reference group;
<sup>b</sup> adjusted for age, sex, household size, migrant background, urban–rural residence, region (east/west), date of participation, and dummies for missing values (separate models for education and income);
<sup>c</sup> self-reported positive throat swab test before study participation.

## DISCUSSION

This is the first German nationwide study of SEP differences in SARS-CoV-2 infections based on individual-level data from the pandemic. The results indicate a higher risk of infection among low-qualified adults than among their highly qualified counterparts. Moreover, the findings suggest socioeconomic differences with respect to the detection of SARS-CoV-2 infections during the pandemic period, with undetected infections being least common among the highly educated.

The strengths of this study include the large Germany-wide sample and its prospective cohort design with comprehensive laboratory confirmation of past and current infections. This design enabled the detection of not only known infections but also infections that had previously gone undetected, for example in asymptomatic cases. Moreover, adding PCR to serological testing was especially relevant in this study because the specimens were collected during the ongoing second wave of SARS-CoV-2 infections in Germany, in which seroconversion from infections in this wave was still in progress. In the analysis, completely non-random non-response could be counteracted by complex weighting, which enabled extrapolation to Germany’s adult population in private households. An important limitation to be considered is that the sample was restricted to residents in private households. Institutionalised people, such as nursing home residents or people living in shared accommodations for homeless people, migrant workers or asylum seekers, are not represented by the sample. Assuming increased infection risks in these groups, this study may have underestimated the nationwide prevalence of SARS-CoV-2 and socioeconomic differences in infections.

Our findings support evidence from earlier pandemics of viral respiratory pathogens, such as influenza, suggesting higher levels of viral exposure in low-SEP settings [6]. Consistent with our findings, higher levels of SARS-CoV-2 infections in low-educated individuals have been found in the UK Biobank [9]. However, individual-level nationwide findings on SEP differences in SARS-CoV-2 infections during the pandemic are still scarce and sometimes contrary [10]. Important mediators in the relationship between education and infections may be occupational working conditions. People in medium-to low-skilled occupations such as nursing, retailing, food production or logistics, had few opportunities to reduce occupational contact and mobility by working remotely during the pandemic, and are associated with more contact and proximity with others.

Our findings indicate an increased risk of SARS-CoV-2 infection in educationally disadvantaged groups and suggest higher detection of infections among highly educated adults. Infection control strategies should provide universal access to testing that is independent of socioeconomic background from early on in order to trace more infections and reduce transmissions from undetected cases. To promote health equity in the pandemic and beyond, social determinants should be given more recognition in infection protection and pandemic planning.

## Data Availability

The data cannot be made publicly available because informed consent from participants did not cover public deposition of data. The minimal dataset underlying the analysis in this article is archived in the Research Data Centre at the Robert Koch Institute in Berlin and can be accessed on site upon reasonable request. The Research Data Centre is accredited by the German Data Forum according to uniform and transparent standards. Requests should be submitted to fdz[at]rki.de.

## DECLARATIONS

## Acknowledgements

We would like to thank all our colleagues at the Robert Koch Institute and the Socio-Economic Panel at the German Institute for Economic Research for their support and cooperation. Special thanks are due to Angelika Schaffrath Rosario for statistical advice and Hans Walter Steinhauer for his contribution to weighting the sample. Our sincere thanks for the laboratory analyses are due in particular to Janine Michel, Martin Schlaud, Silke Stahlberg, Antje Kneuer and Andreas Nitsche. We would also like to thank the employees of Kantar GmbH who contributed to the field work and data collection. We sincerely thank all study participants for their willingness to participate.

## Availability of data and material

The data cannot be made publicly available because informed consent from participants did not cover public deposition of data. The minimal dataset underlying the analysis in this article is archived in the Research Data Centre at the Robert Koch Institute in Berlin and can be accessed on site upon reasonable request. The Research Data Centre is accredited by the German Data Forum according to uniform and transparent standards. Requests should be submitted to.

## Ethical approval

The study was approved by the Ethics Committee of the Berlin Chamber of Physicians (Eth-33/20) in compliance with the Declaration of Helsinki.

## Consent to participate

Written informed consent was obtained from all participants.

## REFERENCES

1. Böhmer MM, Buchholz U, Corman VM, Hoch M, Katz K, Marosevic DV, et al. Investigation of a COVID-19 outbreak in Germany resulting from a single travel-associated primary case: a case series. Lancet Infect Dis. 2020;20(8):920-8. doi:https://doi.org/10.1016/S1473-3099(20)30314-5

2. Wachtler B, Michalski N, Nowossadeck E, Diercke M, Wahrendorf M, Santos-Hövener C, et al. Socioeconomic inequalities and COVID-19: a review of the current international literature. Journal of Health Monitoring. 2020;5(S7):3-17. doi:http://dx.doi.org/10.25646/7059

3. Hoebel J, Michalski N, Wachtler B, Diercke M, Neuhauser H, Wieler LH, et al. Socioeconomic differences in the risk of infection during the second SARS-CoV-2 wave in Germany. Dtsch Arztebl Int. 2021;118:269–70. doi:10.3238/arztebl.m2021.0188

4. Blundell R, Costa Dias M, Joyce R, Xu X. COVID-19 and Inequalities. Fiscal Studies. 2020;41(2):291-319. doi:https://doi.org/10.1111/1475-5890.12232

5. Schröder C, Entringer T, Goebel J, Grabka MM, Graeber D, Kroh M, et al. COVID-19 Is Not Affecting All Working People Equally: DIW Berlin 2020.

6. Quinn SC, Kumar S. Health inequalities and infectious disease epidemics: a challenge for global health security. Biosecurity and bioterrorism : biodefense strategy, practice, and science. 2014;12(5):263–73. doi:10.1089/bsp.2014.0032

7. Hoebel J, Busch M, Grabka MM, Zinn S, Allen J, Gößwald A, et al. Seroepidemiological study on the spread of SARS-CoV-2 in Germany: Study protocol of the ‘CORONA-MONITORING bundesweit’ study (RKI-SOEP study). Journal of Health Monitoring. 2021;6(S1):2-16. doi:http://dx.doi.org/10.25646/7853

8. Robert Koch Institute. Application of the Euroimmun anti-SARS-CoV-2-S1-IgG ELISA antibody test to dried blood spots. 2021. doi:10.25646/8671

9. Niedzwiedz CL, O’Donnell CA, Jani BD, Demou E, Ho FK, Celis-Morales C, et al. Ethnic and socioeconomic differences in SARS-CoV-2 infection: prospective cohort study using UK Biobank. BMC Medicine. 2020;18(1):160. doi:10.1186/s12916-020-01640-8

10. Kastytis Š, Kęstutis P, Vytautas K, Ramunė K, Audronė J, Snieguolė K, et al. SARS-CoV-2 Seroprevalence in Lithuania: Results of National Population Survey. Acta medica Lituanica. 2021;28(1). doi:10.15388/Amed.2020.28.1.2

